# The Influence of Transcatheter Aortic Valve Replacement on Left Atrial Mechanics: A Systematic Review and Meta-Analysis

**DOI:** 10.1101/2023.10.31.23297885

**Authors:** Thomas Meredith, Lauren Brown, Farhan Mohammed, Amy Pomeroy, David Roy, David WM Muller, Christopher Hayward, Michael Feneley, Mayooran Namasivayam

**Author notes:** **Address for correspondence:** A/Prof Mayooran Namasivayam, MBBS, PhD, FRACP, FASE, Department of Cardiology, Level 4, Xavier Building, St Vincent’s Hospital, 438 Victoria Street, Darlinghurst, New South Wales, Australia, 2010. **Disclosures:** A/Prof Namasivayam’s laboratory has received an Nvidia Corporation Academic Hardware Grant. The remaining authors have nothing to disclosure.

## Abstract

**Background:** The morphology and function of the left atrium (LA) are intimately tied to left ventricular loading conditions. Data pertaining to the effect of transcatheter aortic valve replacement (TAVR) on LA function and geometry are scarce.

**Objectives:** To quantify associations between TAVR and LA remodelling by pooling available data from published observational studies.

**Methods:** A systematic review and meta-analysis were performed. Eligible studies needed to report serial LA STE data, before and after TAVR. Other outcome data included LA area and indexed volume (LAVi) and standard chamber measurements. Outcomes were stratified by timing of follow-up echocardiography: early (<6mo) or late (≥6mo).

**Results:** Twelve studies were included, comprising 1,066 patients. The mean overall reduction in LAVi was 2.72mls/m^2^ following TAVI (95% CI 1.37-4.06, p <0.01, low heterogeneity: I^2^ = 0%). LA reservoir function improved overall by a mean difference of 3.71% (95% CI 1.82-5.6, p<0.01), though there was significant heterogeneity within the pooled studies (I^2^ = 87.3%). Significant improvement in reservoir strain was seen in both early follow up (MD 3.1%, p<0.01) and late follow up studies (MD 4.48%, p=0.03), but heterogeneity remained high (I^2^ = 65.23% and 94.4%, respectively). Six studies reported change in LA booster/contractile function, which recovered in the early follow-up studies (MD 2.26, p<0.01), but not in the late group (MD 1.41, p=0.05). Pooled improvement in LA booster function was 1.96% (95% CI 1.11-2.8, p<0.01, low heterogeneity: I^2^ = 0%).

**Conclusion:** The afterload reduction afforded by TAVR is associated with significant haemodynamic and morphological up-stream LA changes.

**Condensed Abstract:** The morphology and function of the left atrium (LA) are intimately tied to left ventricular loading conditions. LA function, measured with speckle-tracking echocardiography (STE), has been demonstrated to provide independent prognostic information for a range of cardiomyopathic states and valvular diseases. We sought to better understand the effect of transcatheter aortic valve replacement (TAVR) on LA function and geometry by performing a systematic review and meta-analysis. Key findings are that, following TAVR, the left atrium negatively remodels (reduces in size), and this is associated with improved distensibility, as quantified by an improvement in reservoir function.

## Introduction

Progressive aortic stenosis is associated with hypertrophic remodelling of the left ventricle, ultimately leading to reactive fibrosis and impaired ventricular function, which is associated with poor prognosis.^1^ Advances in echocardiographic techniques, specifically speckle tracking echocardiography (STE), have facilitated more granular assessment of tissue deformation and mechanical cardiac function. STE reveals early myocardial dysfunction in patients with significant AS prior to symptom development or overt deterioration in systolic function, quantified by the left ventricular ejection fraction (LVEF).^2^ Owing to this robust sensitivity to subtle impairment in ventricular function, strain imaging also provides important prognostic information.^3^ Left atrial (LA) strain imaging can be employed to quantify the phases of left atrial function during the cardiac cycle. LA mechanical function can be divided into three key phases: 1) reservoir phase - collection of pulmonary venous flow during ventricular systole; 2) conduit phase – passive transit of blood to the left ventricle during early ventricular diastole; and 3) contractile/booster phase – active contraction of the atrium during late ventricular diastole.^4^ The triphasic strain curve produced by STE during the cardiac cycle quantifies left atrial distensibility and contractility during these phases. LA mechanical function, measured with STE, has demonstrated independent prognostic discriminatory power for a range of cardiomyopathic states and valvular diseases, including aortic stenosis.^5–7^ Impaired LA function also predicts major adverse cardiac events.^8^ However, the potential reversibility of left atrial mechanical dysfunction following transcatheter aortic valve replacement (TAVR), and the potential prognostic implications are scarcely reported. As such, we sought to quantify these associations by pooling available data from published observational studies.

## Methods

This systematic review was performed according to the Preferred Reporting Items for Systematic Reviews and Meta-Analyses (PRISMA) guidelines.

### Eligibility

To be eligible for inclusion, studies needed to report serial left atrial strain measurements, as quantified by speckle-tracking echocardiography (STE), both before and after transcatheter aortic valve implantation. Prospective and retrospective studies were included.

### Search Strategy

A systematic search of PubMed, Embase and Web of Science was conducted using combinations of the following MeSH and keyword terms: transcatheter aortic valve implantation; TAVI; transcatheter aortic valve replacement; TAVR; left atrial strain; atrial strain; strain. The full search strategy is available in Supplementary Table S1. The databases were queried from their inception until October 2023. To ensure comprehensive capture, an additional manual reference check of pertinent literature including recent review articles was performed.

### Data Extraction and Management

A standardised, pre-piloted form was used to extract data from included studies. Two reviewers (T.M. and L.B.) independently extracted data. Discrepancies were discussed following cross-check. Extracted data included information pertaining to study type, methodology, strain software, and population characteristics. Echocardiographic variables of interest were the aortic valve area (AVA), mean aortic valve gradient, left ventricular ejection fraction (LVEF), left atrial area (LAA), left atrial volume indexed to body surface area (LAVi), and left atrial function, quantified by speckle tracking echocardiography (STE).

### Assessment of Bias

Studies were assessed for risk of bias and methodological quality using the Newcastle-Ottowa tool for assessing risk of bias in cohort studies.^9^ Included studies were rated on 3 different domains, including the selection of the study groups, the comparability of the groups, and the ascertainment of the outcome. The quality score ranges from 0 to 9 points, where 1-3, 4-6, and 7-9 points are reflecting a high, intermediate, and low risk of bias respectively.

### Data Analysis

Analyses were performed using R software (R Foundation for Statistical Computing, Vienna, Austria). Change in left atrial strain parameters from baseline were stratified by duration of follow-up (<6 months and ≥6 months). Where continuous data were reported as median and interquartile range, means were estimated according to the method described by Luo et al^10^, and standard deviations according to the method described by Wan et al. (2014).^11^ Due to the paucity of studies and heterogeneity of outcome reporting, some results are presented qualitatively with relevant figures. Where data were sufficiently homogeneous to permit pooling and meta-analysis, differences were expressed as the mean difference (MD) with 95% CI for continuous outcomes. Studies reporting peak atrial longitudinal strain (PALS) according to the recommendations of the EACVI/ASE/Industry Task Force^12^, which corresponds to reservoir function, were pooled with studies specifically reporting reservoir function. The Hedges random-effects model was used, which has utility in correcting for bias in small sample sizes. Heterogeneity was assessed using the I^2^ statistic, with an I^2^ >50% indicating significant heterogeneity. Potential bias was also visualized using Funnel plots. Where significant heterogeneity existed, meta-regression was performed. A 2-tailed value of P = 0.05 was used to claim statistical significance.

## Results

A total of 697 studies were identified from the search. Following duplicate removal and initial screening for eligibility, 61 full-text studies were retrieved for assessment. 49 studies were excluded, leaving 12 studies for final inclusion (Figure 1).

**Figure 1:**
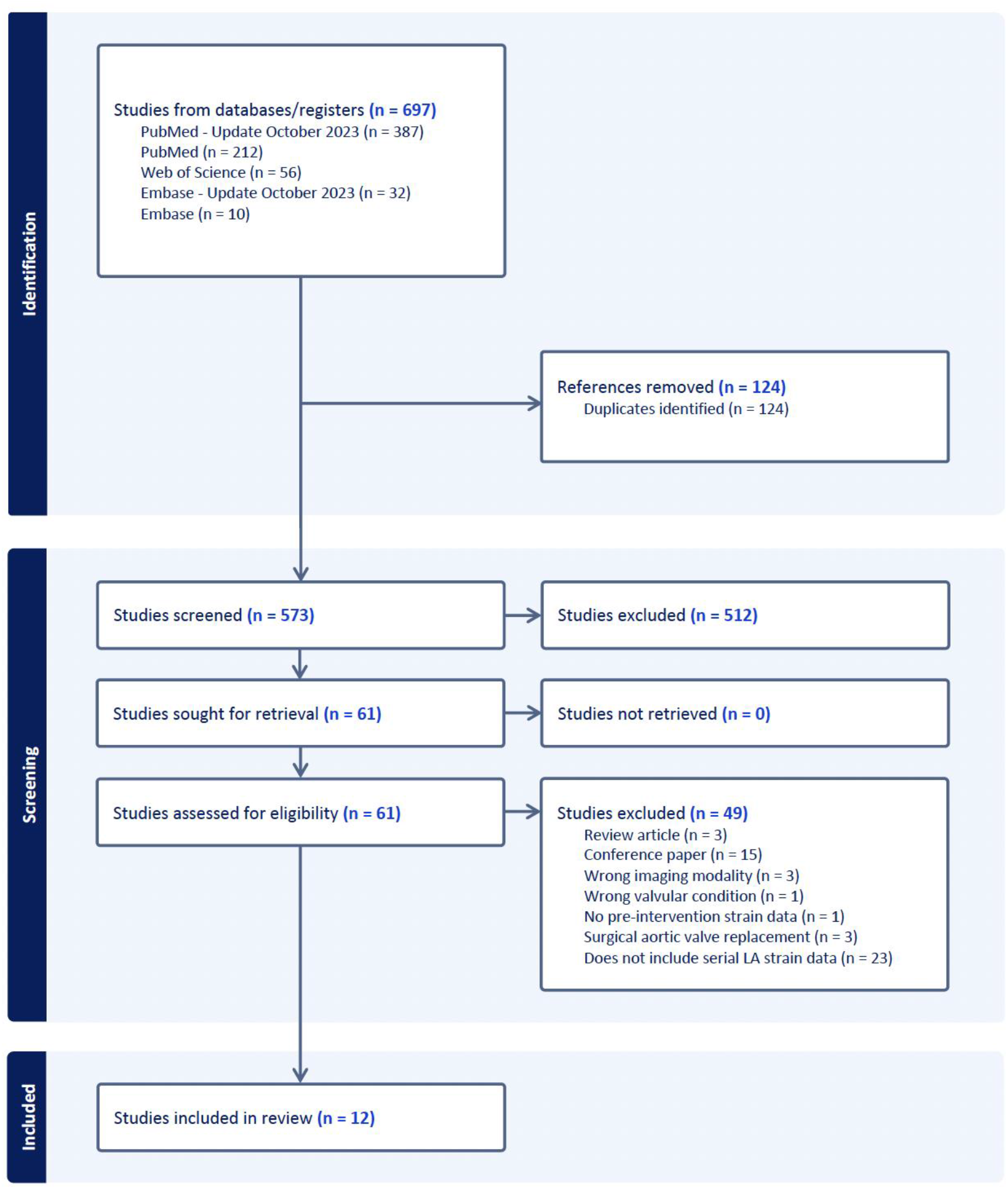
PRISMA flowchart for record screening and assessment. Caption: Initial search yielded 222 studies, from which 11 were eligible for inclusion.

### Baseline Characteristics

Characteristics of the included studies are outlined in Table 1. The pooled cohort totalled 1,066 patients. The average age of participants was 80.8 years. The median LVEF and AVA were 53.7% (IQR 53-58.8) and 0.71cm^2^ (IQR 0.69-0.73), respectively. Seven of the 12 studies reported early follow up data, and most participants underwent TAVR with a self-expanding valve (SEV), compared to a balloon expandable valve (BEV). Six studies utilised EchoPAC strain software, ahead of TomTec and QLab (used in two studies each). All studies demonstrated intermediate risk of bias.

**Table 1:**
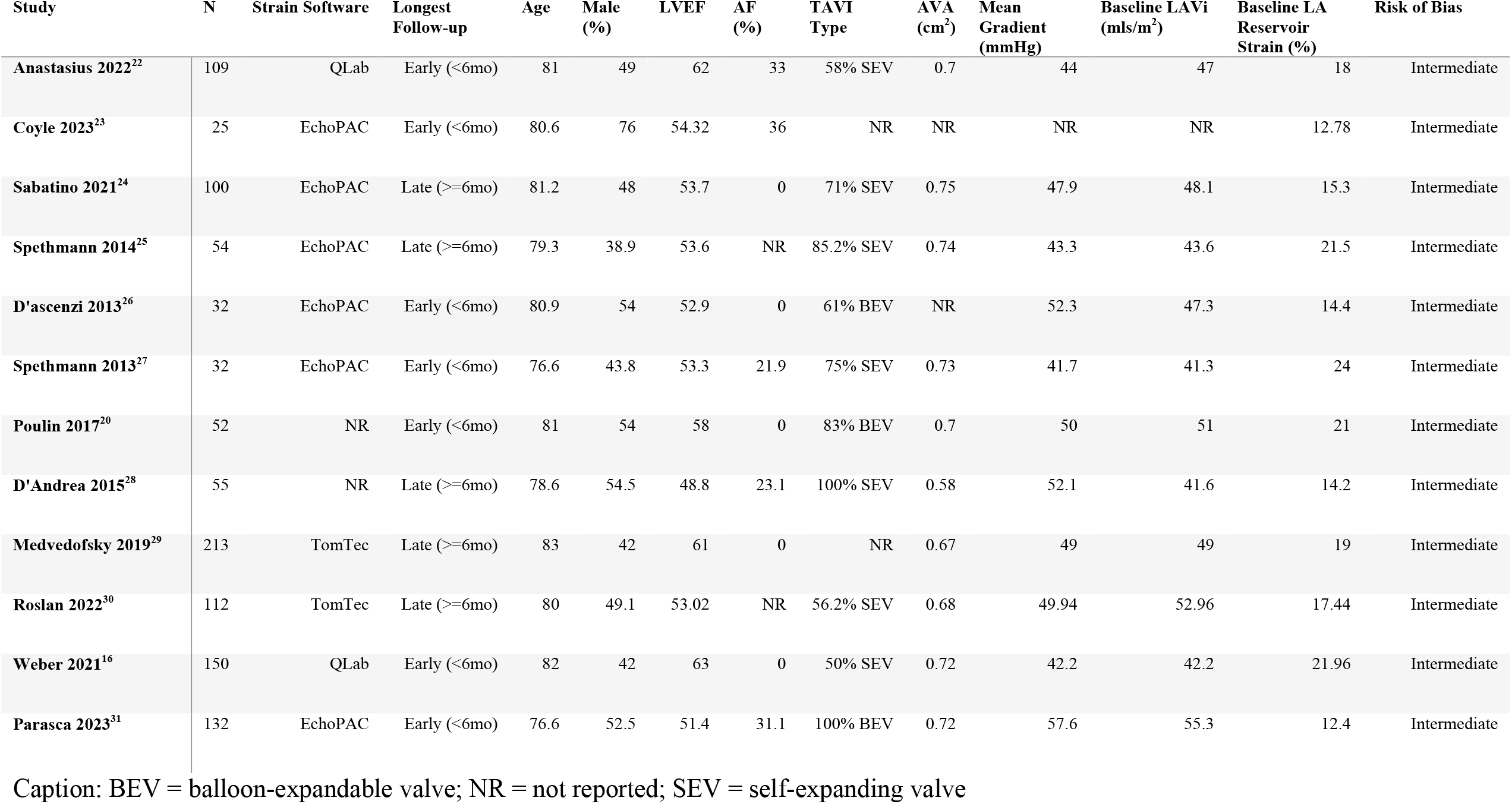
Characteristics of included studies.

### Left Atrial Geometry

Eleven of the twelve studies reported change in LAVi (Figure 2). The mean overall reduction in LAVi was 2.72mls/m^2^ following TAVI (95% CI 1.37-4.06, p <0.01, low heterogeneity: I^2^ = 0%). When stratified by duration of follow up (early or late), a significant change was seen in both early and late follow up groups (Supplementary Figures S1 and S2).

**Figure 2:**
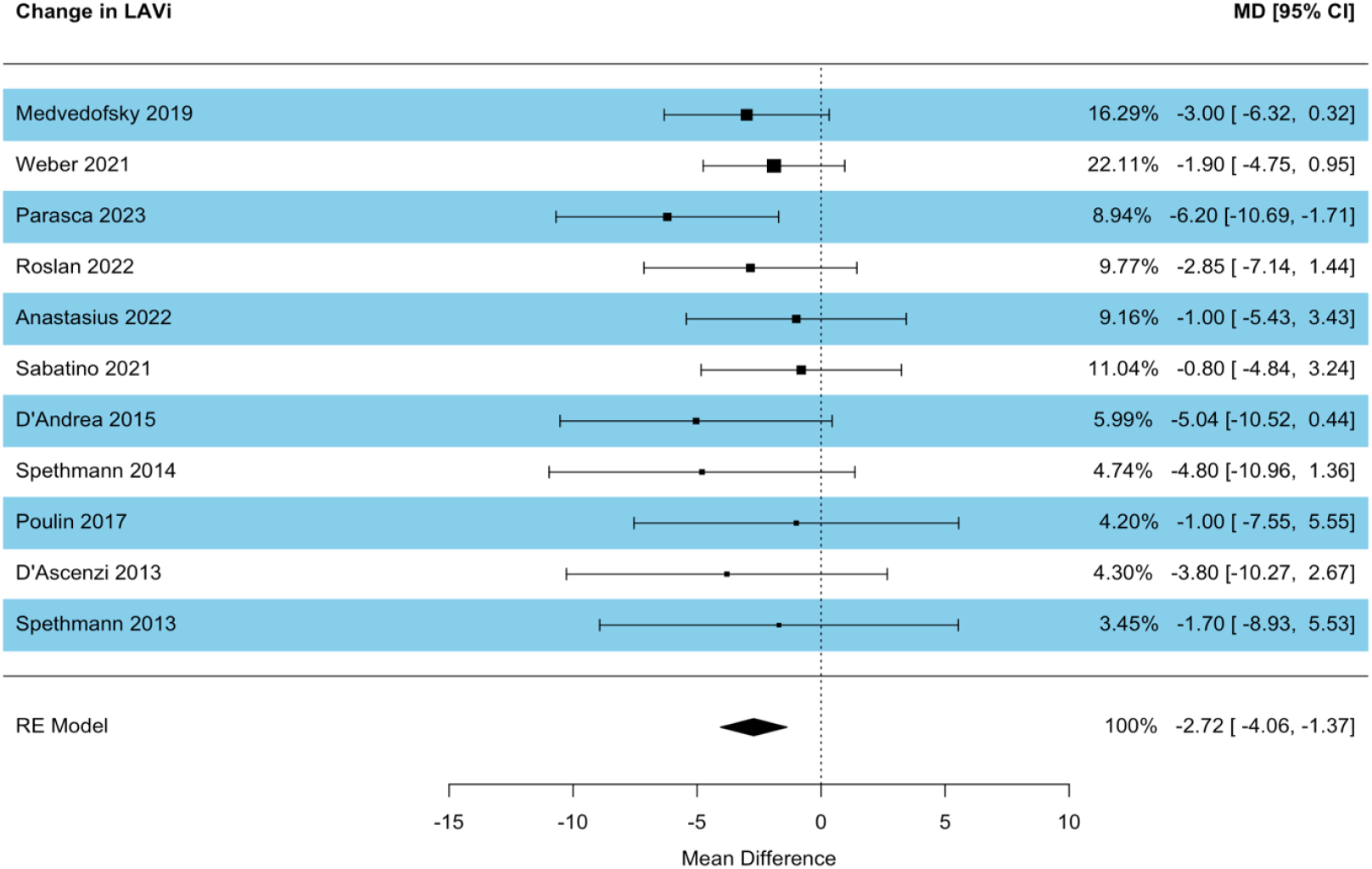
Forest plot of studies reporting change in LAVi (mls/m^2^). Caption: Studies are listed in descending order of sample size.

### Left Atrial Function

All studies reported reservoir indices, albeit six studies referred to this as PALS, and six specifically as reservoir strain. LA reservoir function improved overall by a mean difference of 3.71% (Figure 3, 95% CI 1.82-5.6, p<0.01), though there was significant heterogeneity within the pooled studies (I^2^ = 87.3%). Significant improvement in reservoir strain was seen in both early follow up (MD 3.1%, p<0.01) and late follow up studies (MD 4.48%, p=0.03), but heterogeneity remained high (I^2^ = 65.23% and 94.4%, respectively). Funnel plot visualisation demonstrated some asymmetry, though this was not statistically significant (p = 0.12, Supplementary Figure S3). Meta-regression was therefore performed to test the influence of the following study characteristics: baseline LVEF, proportion of study participants with atrial fibrillation (AF), mean study age, and strain software used. Using a Knapp and Hartung adjustment (given mixed continuous and categorical variables), meta-regression did not reveal statistically significant influence of these variables (F statistic = 1.12, p = 0.55) with significant residual heterogeneity (I^2^ = 84.5%). Although not significant, there was a trend towards greater improvement in reservoir function with lower baseline LVEF (Supplementary Figure S4).

**Figure 3:**
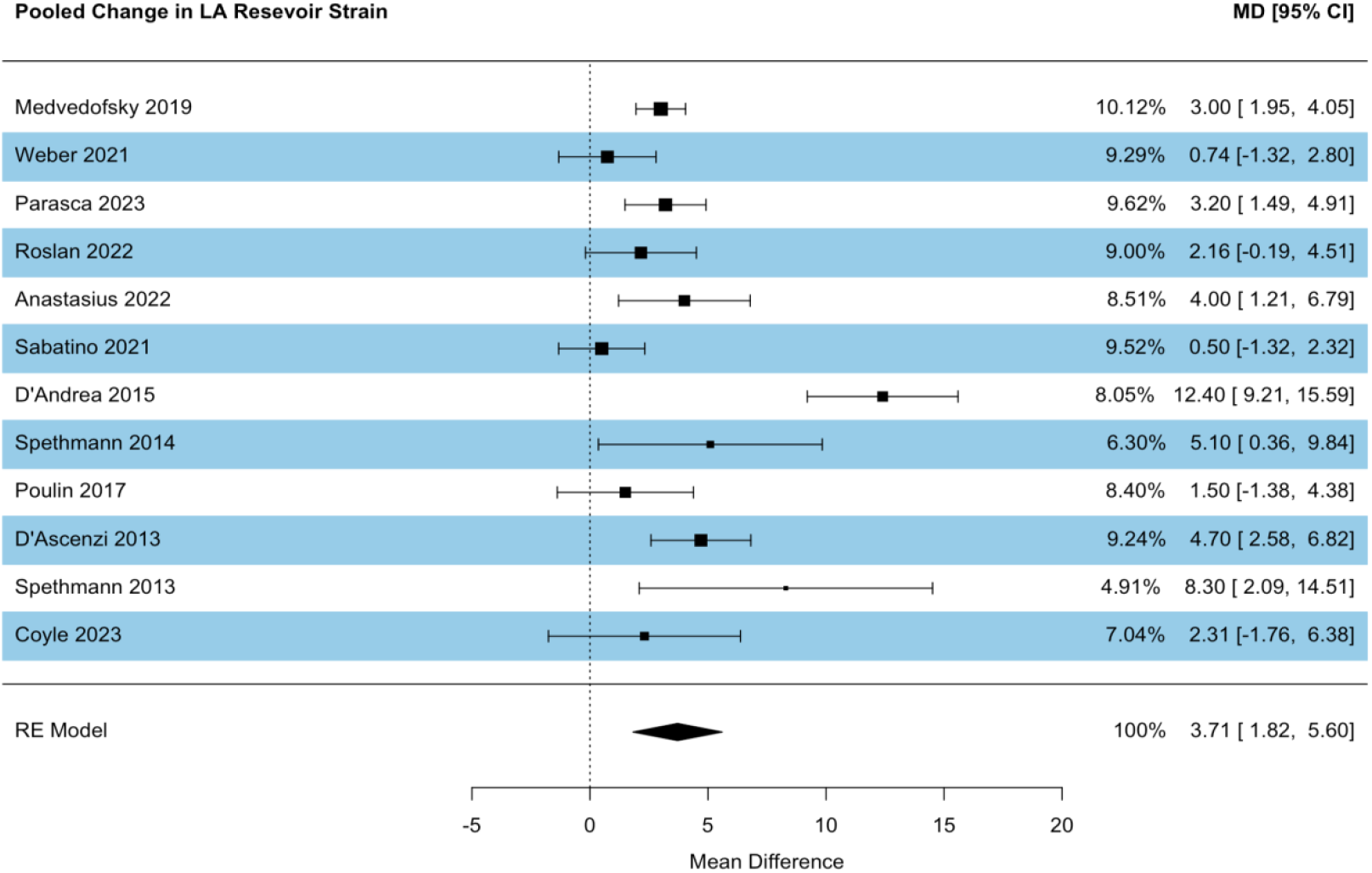
Forest plot of studies reporting change in LA reservoir strain. Caption: Studies are listed in descending order of sample size.

Six studies reported change in LA booster/contractile function, which recovered in the early follow-up studies (MD 2.26, p<0.01), but not in the late group (MD 1.41, p=0.05). Pooled improvement in LA booster function was 1.96% (Figure 4, 95% CI 1.11-2.8, p<0.01, low heterogeneity: I^2^ = 0%). Five studies reported conduit indices. There was a high level of heterogeneity (I2 = 90.1%). The mean improvement in conduit strain was 2.23%, but this was not statistically significant (Figure 5, 95% CI -0.56-5.02, p = 0.12). No significant difference was observed when studies were stratified. Meta-regression was not performed owing to the low number of studies reporting this metric.

**Figure 4:**
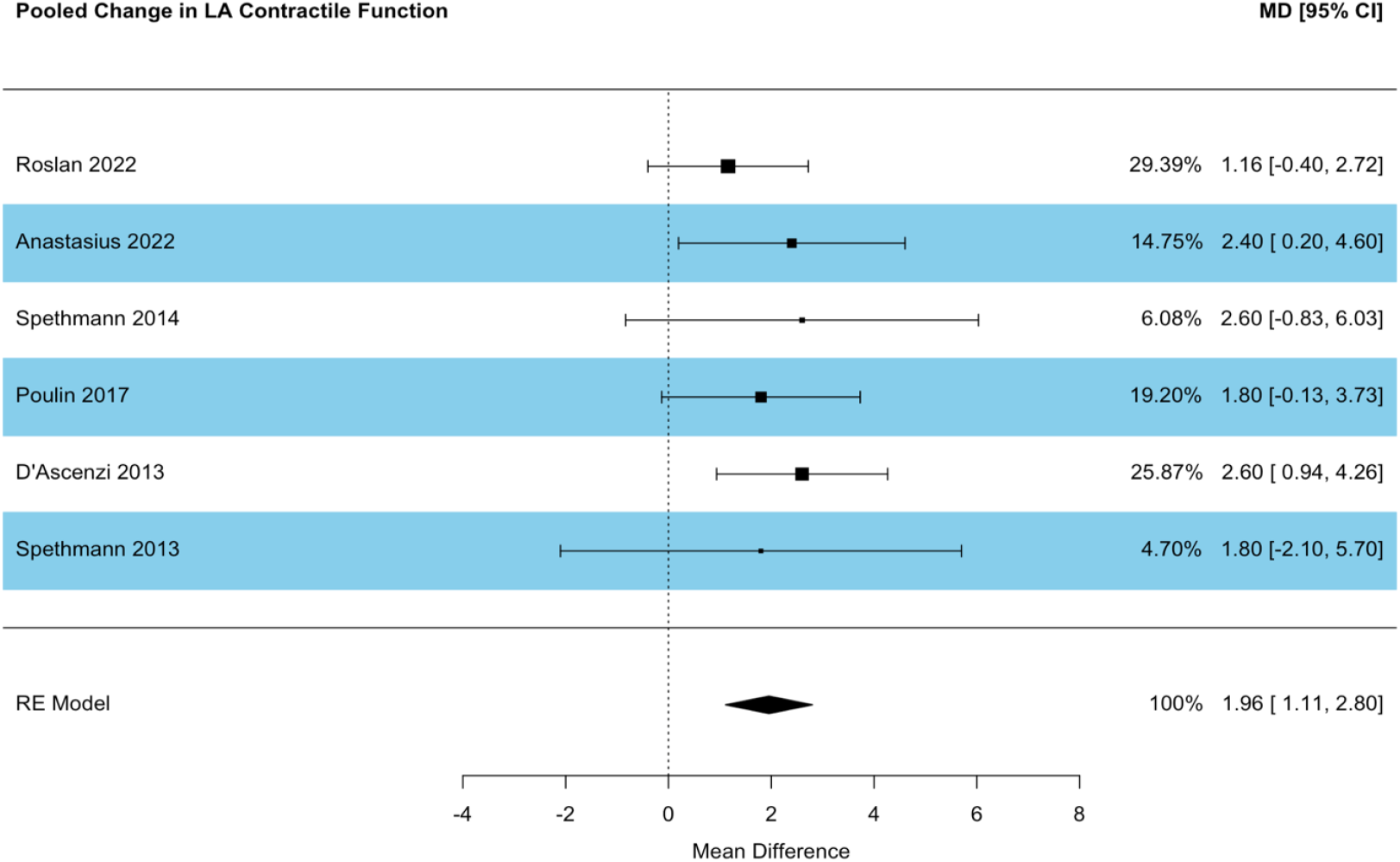
Forest plot of studies reporting change in contractile/booster function. Caption: Studies are listed in descending order of sample size.

**Figure 5:**
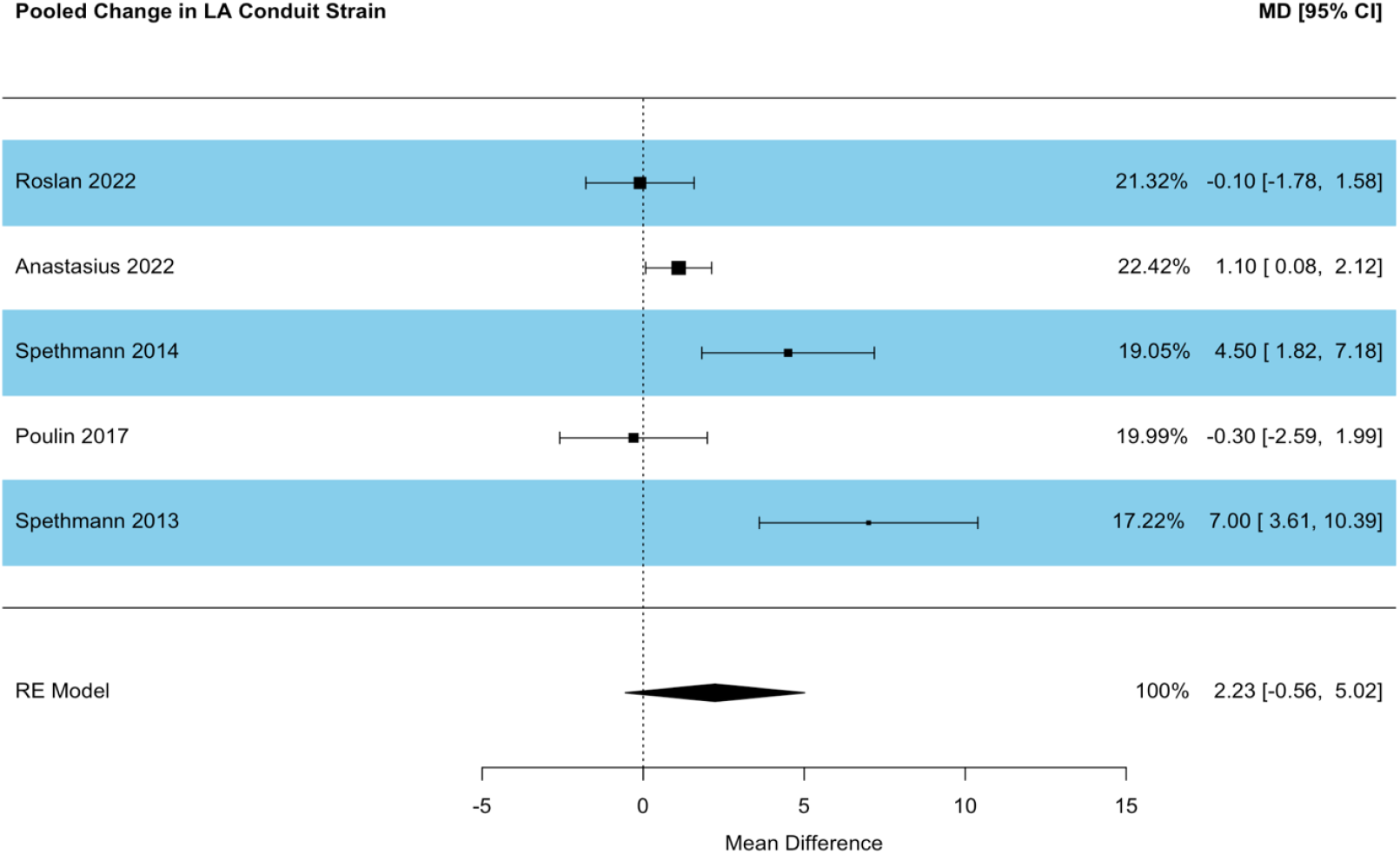
Forest plot of studies reporting change in conduit strain. Caption: Studies are listed in descending order of sample size.

## Discussion

The aim of our study was to quantify improvement, if any, in left atrial mechanical function following TAVR, as quantified by STE. The key findings of this meta-analysis are that, following TAVR, the left atrium negatively remodels (reduces in size), and this is associated with improved distensibility, as quantified by an improvement in reservoir function. It is interesting to observe that in all included studies, the mean baseline reservoir strain was impaired by current consensus criteria (<39.4%), despite generally preserved LVEF.^13^ Moreover, the mean LAVi fell into the severe category (≥40 mls/m^2^), in all 10 studies reporting this metric. Left atrial dysfunction is clearly pervasive within patients with severe aortic stenosis, irrespective of the underlying rhythm. Overall, improvement in LAVi equated to a downgrade in at most one severity category (severe to moderate) in only three studies, remaining within the severe range in the other studies. Despite this, our meta-analysis revealed an overall improvement, but not normalisation, in LA reservoir and contractile function following TAVR.

The reversal of left ventricular hypertrophy following aortic valve replacement, and the prognostic implications, have been studied exhaustively.^14^ However, cardiac adaptation to aortic stenosis is non-uniform, not all patients demonstrate reactive hypertrophy to the chronic pressure load of aortic stenosis, and not all patients favourably remodel following treatment.^15^ Nor is pathological cardiac change confined to the valve or left ventricle – important prognostic information is contained within the left atrium. Both left atrial reverse-remodelling and improvement in left atrial strain indices following TAVR have shown association with improved survival, albeit in a small cohort.^16^ It is also well known that dilation of the left atrium and impairment in mechanical function are associated with increased risk of stroke and stroke related to incident atrial fibrillation.^17, 18^ It has recently been reported that LA strain and strain rate are also predictive of new onset AF following AVR, both transcatheter and surgical.^19, 20^ Poulin et al. found that LA early diastolic strain rate, a marker of conduit function, was associated with new AF following TAVR, raising the question of whether improvement in LA strain following TAVR might be associated with stroke risk reduction. This is currently unknown and requires further research.

Several covariates may contribute to heterogeneity of the included studies, including inter-vendor variability, left atrial strain nomenclature, prevalence of atrial fibrillation, and differences in the type of valve prosthesis (which can influence residual transaortic gradient and left ventricular reverse-remodelling). Meta-regression demonstrated that baseline LVEF, AF, age and strain software accounted for some, but not all, of the heterogeneity. Also, measurement of reservoir function is influenced by apical excursion of the mitral annulus in systole. Therefore, any improvement in this phenomenon afforded by TAVR could influence this observation. Ten of the eleven studies reported a normal mean left ventricular function at baseline, so theoretically a significant change in mitral annular excursion following TAVR might not be expected.

Left atrial strain nomenclature warrants highlighting. It is important to note that measurement of LA strain can be referenced to either the onset of the QRS (end of ventricular diastole) or the beginning of the P wave (beginning of atrial systole, late ventricular diastole). This has important implications for the magnitude of deformation measured, and how each phasic component is calculated. For example, peak positive strain corresponds to conduit function when timed to the P wave, and reservoir strain when timed to the QRS. Further confusion arises when left atrial mechanical phases are labelled according to events of the *left ventricle*. For example, Sabatino et al. report ‘left atrial systolic strain’ (LAS), which corresponds to QRS-gated peak positive strain, and therefore the reservoir phase. Weber et al. report LA global peak longitudinal strain (LAGS), corresponding to the same phase. Indeed, six of the eleven studies in this meta-analysis reported peak left atrial longitudinal strain (PALS) or equivalent, and five reported specific reservoir, conduit and contractile strain values. Coyle et al. used PALS nomenclature but did not specify whether P-wave or QRS gating was implemented. The authors infer that PALS reflects reservoir function, suggesting that they used QRS-gating. The remaining studies reporting PALS also utilised QRS referencing. Presently, standardised strain referencing to the QRS, and labelling according to the *left atrial cycle*, is recommended.^12, 21^

### Limitations

Although nine of the eleven studies were prospective in design, most studies did not report several other covariates known to influence left ventricular remodelling following TAVR, such as paravalvular leak. All studies were observational cohort studies, and we did not analyse patient-level data. Nevertheless, the strength of our study lies in the pooling of multiple studies yielding a combined cohort of 1,066 patients, representing the only meta-analysis of left atrial strain dynamics following TAVR to date.

## Conclusion

The left atrium plays a critical role in modulating left ventricular filling and function. STE can be used to quantify LA mechanical function and has demonstrated independent prognostic power across a range of cardiac conditions including aortic stenosis. Hitherto scarcely described is the extent of LA reverse remodelling following TAVR. This meta-analysis of 12 observational studies revealed significant negative left atrial remodelling following TAVR, and an improvement in left atrial mechanics, quantified by STE. The prognostic implications of these findings require further study.

## Data Availability

All data are available upon reasonable request.

## Acknowledgements

The authors used Covidence (Covidence systematic review software, Veritas Health Innovation, Melbourne, Australia. Available at www.covidence.org).

## Abbreviations

AVA: aortic valve area
BEV: balloon expandable valve
LA: Left atrium
LAVi: Left atrial volume index
LV: Left ventricle
LVEF: left ventricular ejection fraction
SEV: self-expanding valve
STE: speckle tracking echocardiography
TAVR: transcatheter aortic valve replacement

## Figure Legends

**Central Illustration: The Influence of Transcatheter Aortic Valve Replacement on Left Atrial Mechanics: A Systematic Review and Meta-Analysis**

Caption: LA = left atrium; STE = speckle tracking echocardiography; TAVR = transcatheter aortic valve replacement

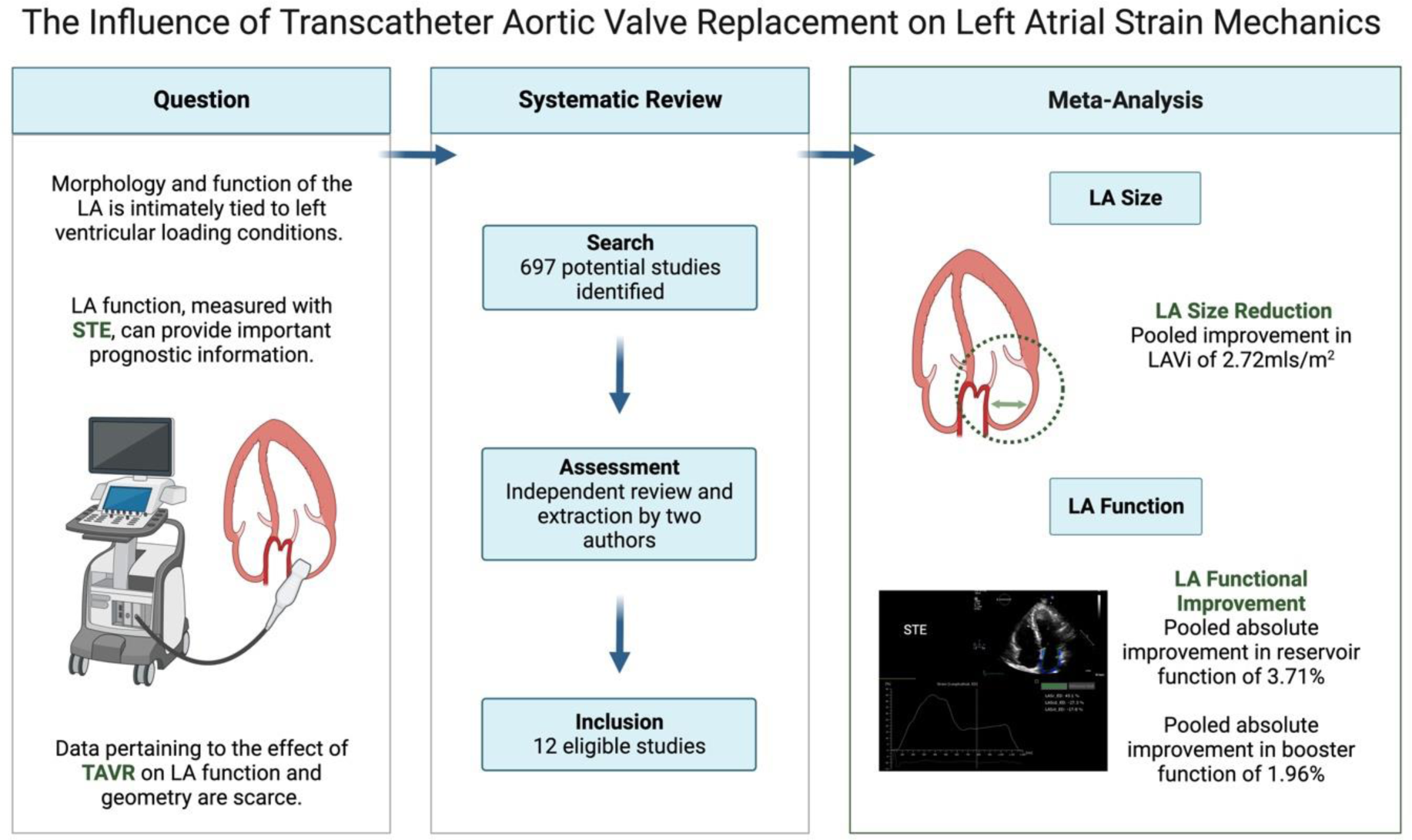

